# Influence of *interleukin-18* polymorphisms on kidney transplantation outcomes: A meta-analysis

**DOI:** 10.1101/2020.05.27.20101196

**Authors:** Thanee Eiamsitrakoon, Phuntila Tharabenjasin, Noel Pabalan, Rungrawee Mongkolrob, Aporn Bualuang, Adis Tasanarong

## Abstract

**Aim:** Allograft survival post-kidney transplantation (KT) are in large part attributed to genetics, which render the recipient susceptible or protected from allograft rejection. KT studies involving single nucleotide polymorphisms (SNPs) have reported the association of *interleukin-18 (IL-18)* with KT and its role in allograft rejection. However, the reported outcomes been inconsistent, prompting a meta-analysis to obtain more precise estimates.

**Methods:** We posed two hypotheses about the *IL-18* SNPs: their association with KT (H1), and increase or decrease in the risks of allograft rejection (H2). Using standard genetic models, we estimated odds ratios [ORs] and 95% confidence intervals by comparing the *IL-18* genotypes between two groups: (i) patients and controls for H1 (GD: genotype distribution analysis); (ii) rejectors and non-rejectors for H2 (allograft analysis). Multiple comparisons were corrected with the Holm-Bonferroni (HB) test. Subgrouping was ethnicity-based (Asians and Caucasians). Heterogeneity was outlier-treated and robustness of outcomes was sensitivity-treated.

**Results:** This metaanalysis generated eight significant outcomes, which HB filtered into four core outcomes, found in the dominant/codominant models. Two of the four were in GD, indicating associations of the *IL-18* SNPs with KT (ORs 1.34 to 1.39, 95% CIs 1.13-1.70, *P*_HB_ = .0007-.004). The other two were in allograft analysis indicating reduced risk with HB P-values of .03 for overall (OR 0.74, 95% CI 0.56-0.93) and Asian (OR 0.70, 95% CI 0.53-0.92). In contrast to the protected Asian subgroup, Caucasians showed non-significant increased risk (OR 1.20. 95% CI .82-1.75, *P*^a^ = .35). Sensitivity treatment conferred robustness to all the core outcomes.

**Conclusions:** Overall association of *IL-18* SNPs with KT was significant (up to 1.4-fold) and Asians KT recipients were protected (up to 30%). Enabled by outlier treatment, these findings were supported by non-heterogeneity and robustness. More studies may confirm or modify our findings.

## l. Introduction

The end-stage of renal failure resulting from kidney disease points to kidney transplantation (KT) as the optimal therapeutic choice [1,2]. The transplanted material (allograft) in the recipient is successful only if it is not rejected [3]. Unrejected allografts are expected to perform the functions as normal kidneys. Normal post-KT graft outcomes depend on immunology where variation in immune responses of the recipient is genetically influenced [4]. This variation may help individualize immunosuppressive regimens by identifying alleles that could increase risk or confer protection for immune-mediated complications [5]. Cytokines are potent immunomodulatory molecules that mediate the immune response [6]. Their production has been shown to be genetically controlled and polymorphisms of many cytokine genes affect their transcriptional activities, resulting in individual variations in cytokine production [7]. Of the cytokine-related factors, interleukin-18 (*IL-18*) has been identified as a post-KT biomarker [8]. Single nucleotide polymorphisms (SNPs) have been reported to be associated with post-KT outcomes [9]. Studies of *IL-18* SNP associations with KT outcomes have promoted better understanding of renal disease immunology, providing greater insight into the biology of KT. However, the primary study conclusions have varied in their degree of concurrence. A meta-analysis addressing this variation may yield clearer estimates of the role of *IL-18* SNPs in KT outcomes. In this meta-analysis, we operated on two hypotheses about the *IL-18* SNPs, their association with KT (H1), and increase or decrease in the risks of AR (H2). In H1, we examine genotype distribution (GD) between patients and healthy controls. In H2 allograft analysis, we compare rejector (RJ) with non-rejector (NRJ) patients. Outcomes from this this might provide useful clinical information for the genetics of KT.

## 2. Methods

### Selection of studies

We searched MEDLINE using PubMed, Google Scholar and Science Direct for association studies as of September 24, 2019. The terms used were “*interleukin*”, “*IL-18* ”, “*cytokine”*, “*polymorphism”*, “*allograft*” and *“renal transplantation*” as medical subject heading and text. References cited in the retrieved articles were also screened manually to identify additional eligible studies. In cases of duplicate articles, we selected the one with a later date of publication. Inclusion criteria were (1) case–control design evaluating the association between *IL-18* polymorphisms and KT outcomes. (2) *IL-18* genotype frequencies that compare KT patients and healthy controls, NR and NRJ. (3) Sufficient genotype frequency data to enable calculation of the odds ratios (ORs) and 95% confidence intervals (CIs). Exclusion criteria were (1) not involving renal allografts or post-KT outcomes; (2) reviews; (3) not about the *IL-18* polymorphisms and (4) studies whose genotype or allele frequencies were unusable or absent.

### SNP groupings

The included articles examined two *IL-18* SNPs, rs187238 and rs1946518, each presented with genotype data (Tables S2 and S3). Observed phenotypic associations have been attributed to the proximity of two SNPs [10,11]. NCI LDLINK (https://ldlink.nci.nih.gov/) results shows that the two SNPS are in linkage disequilibrium (LD). LD is the correlation between alleles located near each other [12] which is measured in terms of D' with a value of 1 indicating complete LD [13]. Therefore, *IL-18* SNPs (rs187238 and rs1946518) with D' values of 1.00 in this study (Table S1) were combined in the analysis (Tables S2 and S3). The rationale for combining rests on the assumption that SNPs in LD yield similar associations in the phenotype.

### Data extraction, HWE and methodological quality

Two investigators (TE and NP) independently extracted data and arrived at a consensus. The following information was obtained from each publication: first author’s name, year of the study, country of origin, ethnicity, age of the subjects in years, *IL-18* SNPs (rs number) (Table 1). Sample sizes as well as genotype data between the RJ and NRJ were also extracted along with calculated outcome of the minor allele frequency (maf) (Tables S2 and S3). The Hardy-Weinberg Equilibrium (HWE) was assessed using the application in https://ihg.gsf.de/cgi-bin/hw/hwa1.pl. The Clark-Baudouin (CB) scale was used to evaluate methodological quality of the included studies [14]. CB criteria include *P*-values, statistical power, correction for multiplicity, comparative sample sizes between cases and controls, genotyping methods and the HWE. In this scale, low, moderate and high have scores of < 5, 5-6 and ≥ 7, respectively.

**Table 1.** Characteristics of the included studies in *interleukin-18* associations with kidney transplantation outcomes

| First author | [R] | Year | Country | Ethnicity | Age (years) mean ± SD<br>RJ / NRJ | *IL-18* SNPs | CB |
| --- | --- | --- | --- | --- | --- | --- | --- |
| Kim | 20 | 2008 | Korea | Asian | 33.9 ± 9.4 / 36.1 ± 11.1 | rs187238 | 10 |
| Mittal | 21 | 2011 | India | Asian | 33.2 ± 12.6 / 38.2 ± 11.1 | rs187238, rs1946518 | 6 |
| Kolesar | 22 | 2007 | Czechlovakia | Caucasian | 49.6 (patients) | rs187238, rs1946518 | 7 |
| do Nascimento | 23 | 2014 | Brazil | Caucasian | 33.1 ± 12.4 / 40.5 ± 13.0 | rs187238, rs1946518 | 7 |

[R] Reference; *IL-18*,; SD, standard deviation; RJ, rejector; NRJ, non-rejector; SNP, single nucleotide polymorphism; CB, Clark-Baudouin

### Meta-analysis

We estimated ORs and 95 % CIs using two overall approaches: (i) genotype distribution (GD) between cases and healthy controls and (ii) allograft wherein RJ were compared with NRJ. Thus, both were KT outcomes were analysed separately. Calculated pooled ORs for GD were either higher in patients (hp) or higher in controls (hc); in allograft, they were either increased (in) or decreased (de), indicating risk for rejection. Standard genetic modeling was used, wherein we compared the following, (i) recessive (Rc: *wt-wt* versus *wt-var* + var-var), (ii) dominant (Do: *wt-wt* + *wt-var* versus *var-var*) and (iii) codominant (Co: *wt* versus var) effects. Heterogeneity between studies was estimated with the χ^2^-based Q test [15], with threshold of significance set at *P*^b^ < .10. Heterogeneity was also quantified with the I^2^ statistic which measures variability between studies [16]. Evidence of functional similarities in population features of the studies warranted using the fixed-effects model [17], otherwise the random-effects model [18] was used. Sources of heterogeneity were detected with the Galbraith plot [19] followed by re-analysis (outlier treatment). Of note, outlier treatment dichotomized the comparisons into pre-outlier (PRO) and post-outlier (PSO). Sensitivity analysis, which involves omitting one study at a time and recalculating the pooled OR, was used to test for robustness of the summary effects. The low number of studies precluded assessment of publication bias. Multiple associative outcomes were Holm-Bonferroni (HB) corrected. Data were analysed using Review Manager 5.3 (Cochrane Collaboration, Oxford, England), sigmaplot 11.0 and sigmastat 2.03 (Systat Software, San Jose, CA, USA).

## 3. Results

### Search outcomes and study features

Figure 1 outlines the study selection process in a PRISMA-sanctioned flowchart (Preferred Reporting Items for Systematic Reviews and Meta-Analyses). Initial search resulted in 39 citations, followed by a series of omissions that eventually yielded four articles for inclusion [20-23]. Table 1 shows two Asian [20,21] and two Caucasian [23,22] articles with middle-age profile of the KT subjects (mean ± SD: 37.8 years ± 5.9). Three [23,22,21] of the four included articles examined the two *IL-18* polymorphisms (rs187238 and rs1946518). Methodological quality of the component studies was moderate with a mean ± SD of 6.37 ± 1.24. Tables S2 and S3 show seven studies each for GD and allograft analyses. This metaanalysis followed the PRISMA guidelines (Table S5).

**Figure 1.**
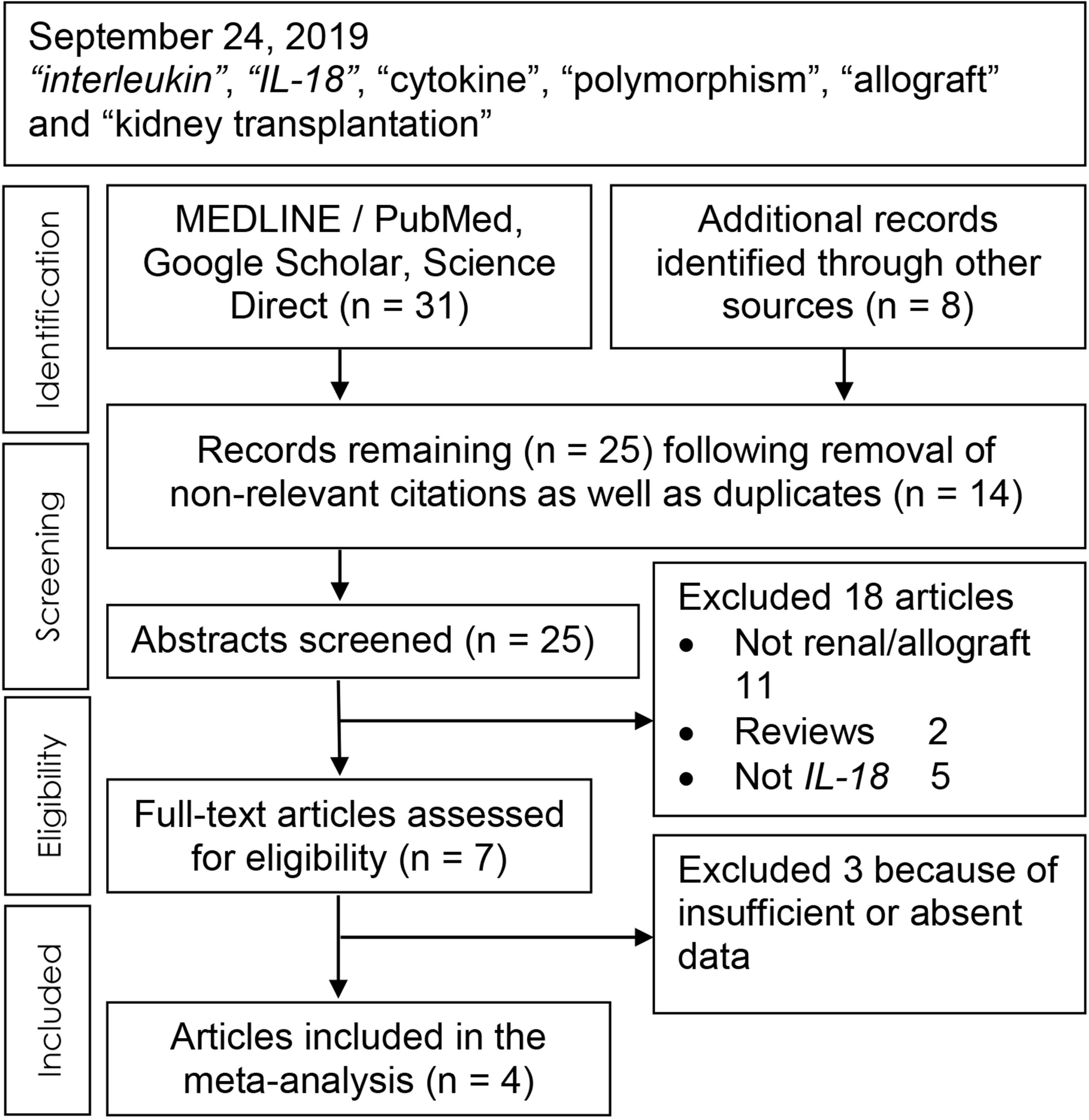
Summary flowchart of literature search

### Meta-analysis outcomes

Table 2 delineates the overall pooled ORs by direction of effect, where GDs were higher in patients (*hp*) (OR > 1.00) but decreased risk (*de*) in the allograft analysis (OR < 1.00). The results generated 27 comparisons (Tables 2-3), eight of which were statistically significant (*P*^a^ < .05). Of the eight, four withstood the HB correction, which were considered the core findings. Of the four, two were in GD showing *hp* effects in the Do/Co models (OR 1.34-1.39, 95% CIs 1.13 to 1.70, *P*_HB_ = .0007-.004). The other two core outcomes (at HB *P*-values of .03) were in allograft analysis indicating reduced risk in the Co model, one in the overall (OR 0.73, 95% CI 0.56-0.93) and the other in Asians (OR 0.70, 95% CI 0.53-0.92). This Asian contrasted with the increased risk outcome in Caucasians (OR 1.20, 95% CI 0.82-1.75, P^a^ = 0.35)

**Table 2.**
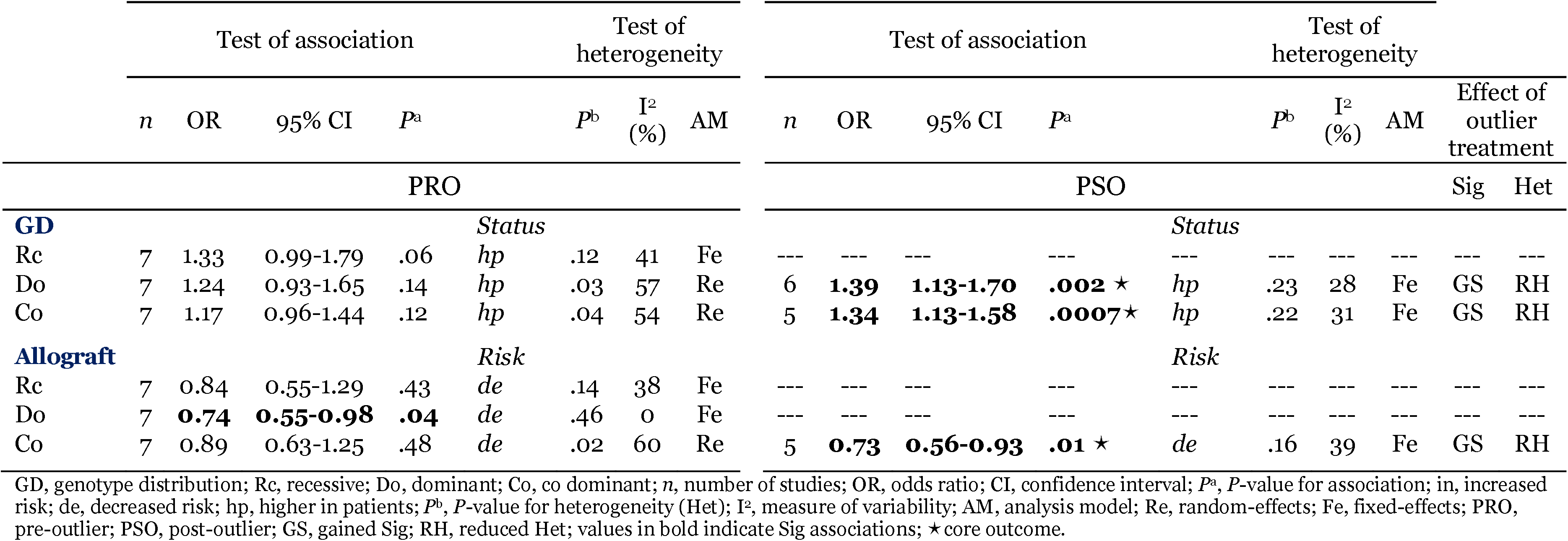
Summary outcomes for associations of *interleukin-18* polymorphisms with kidney transplantation outcomes

**Table 3.**
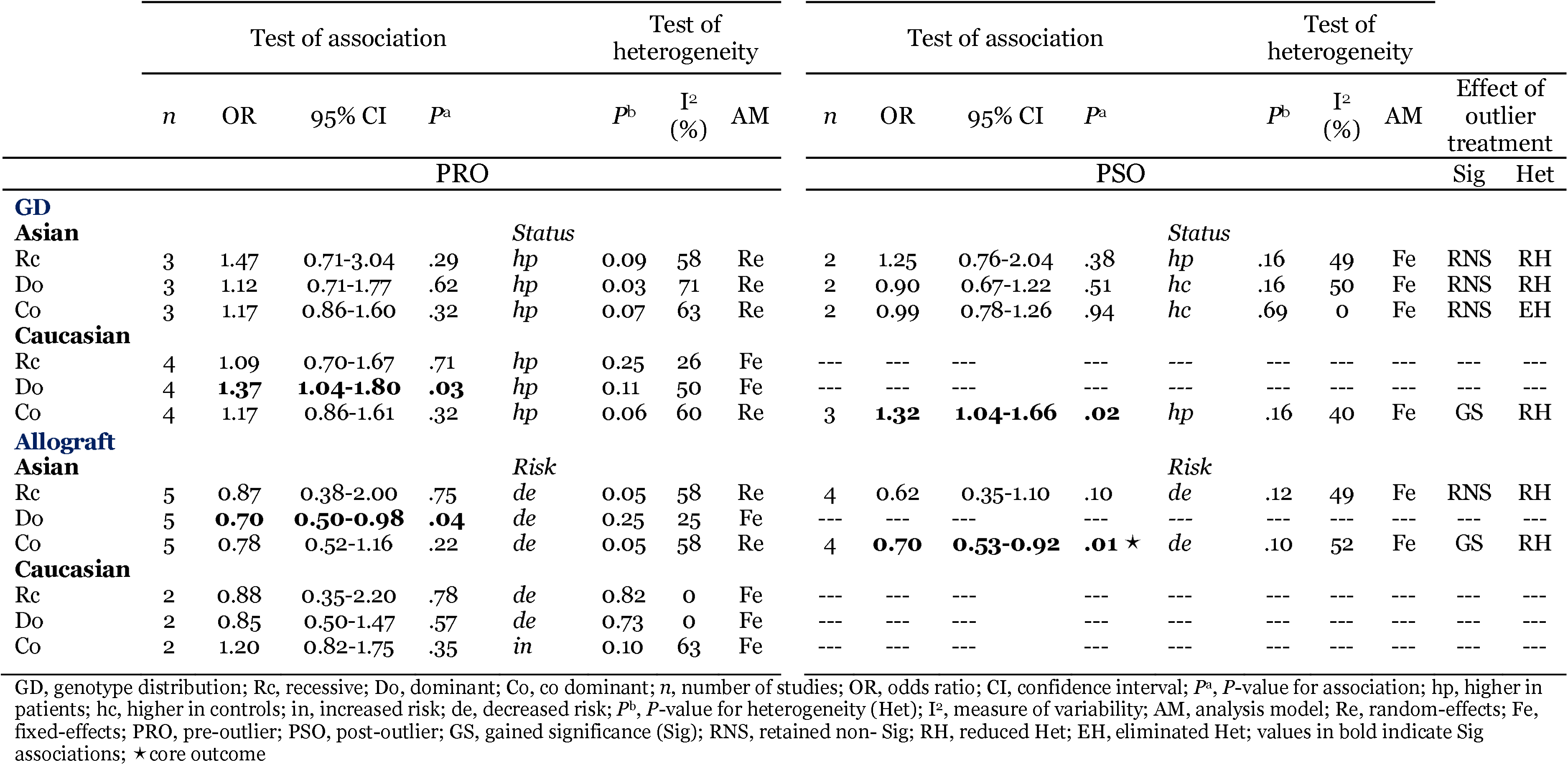
Subgroup outcomes for associations of *interleukin-18* polymorphisms with kidney transplantation outcomes

Of note, all four core outcomes were outlier-derived (PSO). The mechanism of outlier treatment for *IL-18* in the Co model of allograft analysis is visualized in Figures 2-4. Figure 2 shows the PRO forest plot with a non-significant (*P*^a^ = .48) and heterogeneous (*P*^b^ = .02, I^2^ = 60%) pooled effect indicating reduced risk (OR 0.89 95% CI 0.63 to 1.25). The Galbraith plot identified the two studies [22,21] as the sources of heterogeneity (outliers), located above the +2 confidence limit (Figure 3). In Figure 4, the PSO outcome (outliers omitted) shows reduced heterogeneity (*P*^b^ = .16, I^2^ = 39%); reduced risk effect (OR 0.73 95% CI 0.56 to 0.93) and gained significance (*P*^a^ = .01). This operation is numerically summarized in Table 2. Sensitivity treatment deemed the core outcomes to be robust.

**Figure 2.**
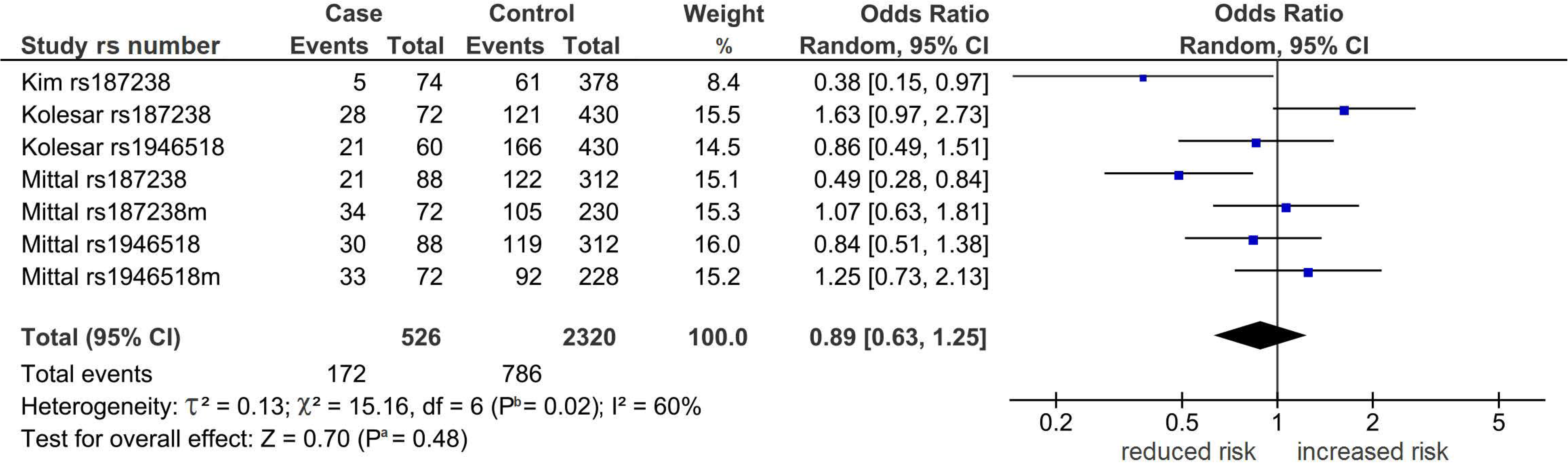
Forest plot outcome in the allograft analysis of the codominant model. Diamond denotes the pooled odds ratio (OR) indicating reduced risk (0.89). Squares indicate the OR in each study. m, match. Horizontal lines on either side of each square represent the 95% confidence intervals (CI). The Z test for overall effect was non-significant (*P*^a^ = .48). The x^2^-test shows the presence of heterogeneity (*P*^b^ = .02, I^2^ = 60%); I^2^, a measure of variability expressed in %

**Figure 3.**
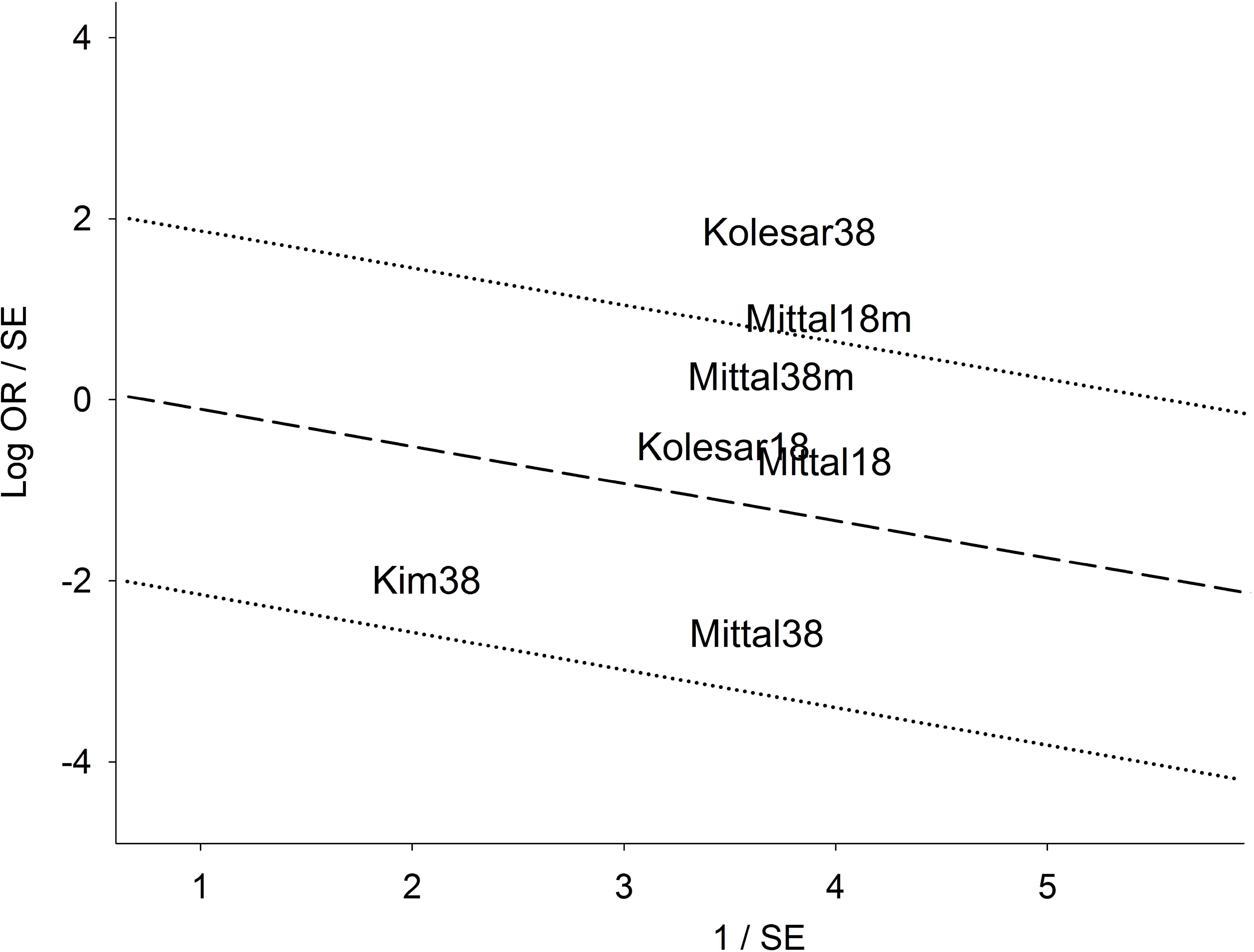
Galbraith plot of the allograft analysis in the codominant model m, match; Log OR, logarithm of standardized odds ratio; SE, standard error. The two studies above the +2 confidence limit are the outliers.

**Figure 4.**
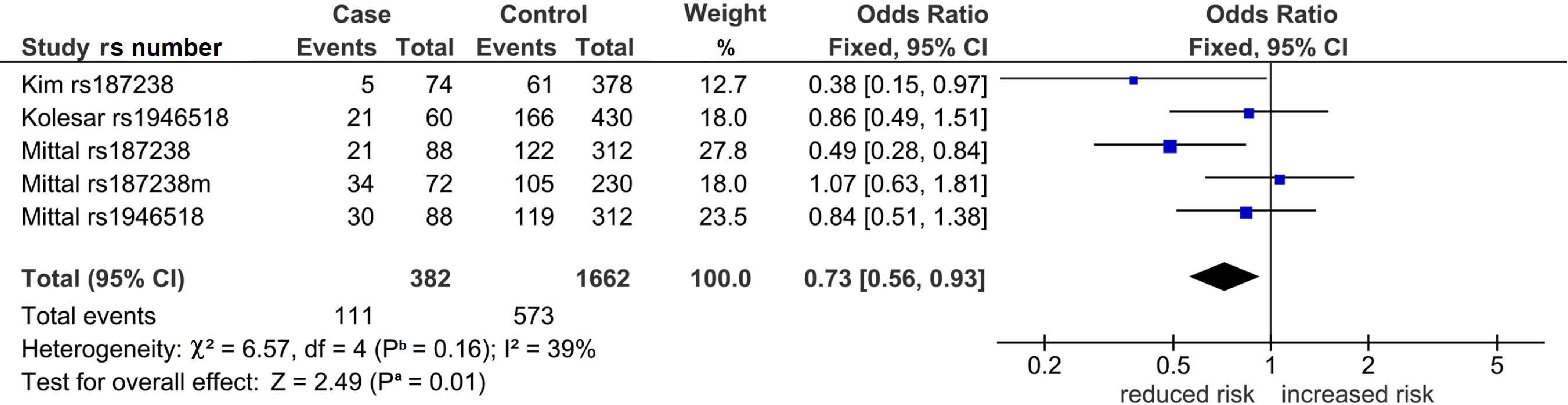
Forest plot outcome of outlier treatment in the allograft analysis of the codominant model. Diamond denotes the pooled odds ratio (OR) reduced risk (0.73). Squares indicate the OR in each study, m, match. Horizontal lines on either side of each square represent the 95% confidence intervals (CI). The Z test for overall effect shows significance (*P*^a^ = .01).The χ^2^-test indicates reduced heterogeneity (*P*^b^ = .16, I^2^ = 39%); I^2^, a measure of variability expressed in %

## 4. Discussion

The main findings of this study showed that *IL-18* SNPs were associated with KT outcomes, more specifically, the allograft analysis indicated reduced risks of rejection. Subgroup analysis identified Asian KT recipients with the *IL-18* SNPs as protected from allograft rejection which contrasted with the increased risk for the Caucasian subgroup. The core status of having withstood HB and robustness of our principal findings underpin the strength of evidence in this study. Furthermore, outlier treatment unraveled significant and non-heterogeneous associations that were not present in the component single-study outcomes. Conflicting outcomes between primary studies may be attributed to their lack of power and small sample sizes. Underpowered outcomes appear to be common in candidate gene studies [24] and are prone to the risk of Type 1 error. In spite of the evidence for associations, the complexity of allograft rejection involves interactions between genetic and non-genetic factors allowing for the possibility of environmental involvement. Gene-gene and gene-environment interactions have been reported to have roles in associations of other polymorphisms with post-KT allograft rejection. One article [21] examined another gene polymorphism (vascular endothelial growth factor (*VEGF)*. All articles acknowledged gene-environment interaction. Addressing gene-gene and gene-environment interactions may help address the pathophysiological significance of *IL-18* in allograft failure post-KT. All the included articles mentioned haplotype analysis with one presenting haplotype data [21]. Focus on *IL-18* haplotypes have been suggested for future association studies [9]. The crucial role of *IL-18* in kidney physiology lies in its involvement in the filtration, integrity and permeability of the glomerular basement membrane [25]. IL-18 expression in the renal epithelium might be important in triggering specific immune response manifested as acute graft rejection [26]. Increased IL-18 production promotes enhanced endothelial permeability and augmented leukocyte migration into the allograft, promoting a clinically recognized rejection episode [21]. A study demonstrated upregulation of IL-18 production in patients with acute rejection of kidney allograft [26]. Moreover, another study found significantly higher levels of IL-18 in culture biopsies from patients with acute rejection in comparison to stable KT patients [27]. Urinary IL-18 has been found to be an early, noninvasive and accurate predictor for dialysis within the first week of KT [28].

## 5. Strengths and limitations

Two strengths of our study were: (i) outlier treatment was key to generating significance and reducing heterogeneity; and (ii) subgrouping identified Asians as significantly protected and Caucasians as non-significantly susceptible to allograft rejection. Limitations include: (i) the component studies were underpowered, however, sample sizes were adequate at the aggregate level with 624 cases/634 controls in GD (Table S2) and 147 cases/674 controls in allograft (Table S3). (ii) Genotype distributions of the control population in some studies deviated from the HWE (Tables S2 and S3) and it might be a source of potential bias in our study.

## 6. Conclusions

To our knowledge, this is the first meta-analysis with evidence that may render *IL-18* useful as a prognostic marker in allograft rejection post-KT. Additional well-designed studies exploring other parameters may confirm or modify our results in this study.

## Data Availability

The data from our study is presented in the Supplementary material

**Abbreviations**
robust (all other significant outcomes: were non-robust)
✓: significant outcome that survived the Bonferroni correction
A: adenine
AM: analysis model
C: cytosine
Co: codominant genetic model
CB: Clark-Baudouin
CC: homozygous genotype
CI: confidence interval
Do: dominant genetic model
de: decreased risk
du: duplicate
EH: eliminated heterogeneity
Fe: fixed-effects
G: guanine
GD: genotype distribution
GS: gained significance
H1: hypothesis 1 (GD analysis)
H2: hypothesis 2 (allograft analysis)
het: heterogeneity
Ho: homozygous genetic model
hc: higher in controls
hp: higher in patients
HWE: Hardy-Weinberg Equilibrium
*IL-18*: *interleukin-18* gene
IL-18: interleukin-18 protein
in: increased risk
KT: kidney transplantation
LD: linkage disequilibrium
Log OR: logarithm of standardized odds ratio
maf: minor allele frequency
*n*: number of studies
NRJ: non-rejector
OR: odds ratio
*P*^a^: P-value for association
*P*^b^: P-value for heterogeneity
PRO: pre-outlier
PSO: post outlier
I^2^: measure of variability
Rc: recessive genetic model
[R]: Reference
Re: random-effects
RH: reduced heterogeneity
RJ: rejector
RNS: retained non-significance
SD: standard deviation
SE: standard error
sig: significant
SNP: single nucleotide polymorphism
*var*: variant
*wt*: wild-type homozygotes
*wt-var*: heterozygote

## Supporting information

Table S1 LD matrix DOCX

Table S2 Quantitative features GD DOCX

Table S3 Quantitative features

Table S4 PRISMA checklist DOCX

